# Predictors of black fungus fear during the COVID-19 pandemic among the Bangladeshi health workers: a cross-sectional study

**DOI:** 10.1101/2021.07.03.21259961

**Authors:** Md. Kamrul Hasan, Humayun Kabir, Mamunur Rahman, Anjan Kumar Roy, Shimpi Akter, Dipak Kumar Mitra

## Abstract

**Background:** The emergence of mucormycosis cases amid the COVID-19 pandemic; fear associated with mucormycosis may turn out to be a terrifying public health issue. This study aimed to assess the association between fear and insomnia status and other predictors of mucormycosis among Bangladeshi healthcare workers.

**Methods:** From 25 May 2021 to 05 June 2021, a cross-sectional study was carried out among healthcare workers. A total of 422 healthcare workers participated in this study. A semi-structured online questionnaire was used for data collection during the COVID-19 pandemic, followed by convenient and snowball sampling methods. A multivariable linear regression model was fitted to assess the association between fear and insomnia status and other predictors of mucormycosis.

**Results:** The results indicated that the respondents with insomnia status had a higher score of mucormycosis fear than not having insomnia (β = 3.91, 95% CI: 2.49, 5.33, *p* <0.001), significantly. Alongside the increased knowledge score of mucormycosis, the average score of fear increased significantly(β = 0.35, 95% CI: 0.20, 0.50, p <0.001). The gender, profession, and death of friends and family members due to COVID-19 significantly affected mucormycosis fear score increment.

**Conclusions:** This is the first study that focused on assessing the association between mucormycosis fear and insomnia status among the health care workers so far. These study findings recommend emphasizing the mental health aspects and ensuring support to the healthcare workers to better tackle the ongoing public health crisis.

## 1. Introduction

Coronavirus disease 2019 (COVID-19), also known as SARS-CoV-2, the ongoing pandemic which was initiated in December 2019 in China [1], has affected Bangladesh adversely like many other countries [2]. Up to 04 June 2021, 0.8 million cases were recorded along with around 7.5 thousand deaths by COVID-19, first diagnosed on 8 March 2020 in Bangladesh according to IEDCR, the country’s epidemiology institute [3], [4]. Mucormycosis is not a newly emerged disease. This disease affected workers in conditions of immunodeficiency. One case worth mentioning is the infection observed in a shipbuilding worker [5]. Mucormycosis, nowadays popularly known as ‘Black fungus’ in the COVID-19 era, is a treacherous fungal infection that recently became a new threat for COVID-19 survived patients who were first diagnosed in Bangladesh on 08 May 2021 [6], [7].

The prevalence of mucormycosis is eighty times more in India during the COVID-19 pandemic [8]. The mucormycosis associated with COVID-19 has affected many countries like Bangladesh, Pakistan, Russia, Iran, and Brazil [9]–[14]. As mucormycosis infection in India associated with COVID-19 has raised at an alarming rate with a high mortality rate of about 50%, it is a concerning issue for Bangladesh being geographically attached with India [15]. The incidence of mucormycosis infection increases more rapidly in the second wave than in the first wave, with at least 14,872 cases on 28 May 2021 [16]. The mucormycosis is caused by the order Mucorales, and mostly it is due to the intrusion of the genre *Rhizopus* and *Mucor* [17], [18]. People are affected by mucormycosis through the contact of the fungal spores, as inhalation of the spore from the environment can cause infection in the lung and sinus [19]. Albeit, without the presence of a biological niche of this fungus, it cannot affect anyone. High-risk mucormycosis infection determinants include immunosuppression, such as hematologic malignancy, hematopoietic stem cell transplantation (HSCT), solid organ transplantation (SOT), and diabetes mellitus [20]. Underlying patients’ disease conditions are considered a major factor for getting a primary infection of mucormycosis [21]–[23]. Nowadays, COVID-19 associated mucormycosis infection is mostly rhino orbital cerebral type infection which is clinically manifested around the nose, eye, and brain [24]. The use of corticosteroid therapy for managing COVID-19 patients and diabetes mellitus is currently indicated as a risk factor for mucormycosis infection [25]. A study found that patient suffering from COVID-19 has been affected with pulmonary mucormycosis primarily due to an immunocompromised state [26]. The mucormycosis pathogenesis includes vessel thrombosis and subsequent tissue necrosis, which causes mortality around 54% [21], [27]. To prevent mucormycosis infection during COVID-19, avoiding the risk factors is the only option. Though the COVID-19 vaccination has started, there is no vaccine currently available for mucormycosis. Therefore, early diagnosis and treatment intervention is essential to treat COVID-19 induced mucormycosis infection effectively.

As front liners, the health care workers are an integral part of the health system fighting against the COVID-19 and serious mucormycosis infection threat [28]. A study found that inadequate hospital infection prevention strategies, workplace safety, occupational injury prevention and poor control of policies were the reasons for fear among the health care workers during any emerging outbreak [29], [30]. In addition, safety protocol for Bangladeshi health workers was found to be insufficient [31]. Therefore, fear and insomnia might be natural phenomena for the Bangladeshi healthcare workers to simultaneously tackle the COVID-19 pandemic and upheaval of mucormycosis. However, mitigating the fear, insomnia, and proper knowledge about the disease and initiatives may guide the health care workers of Bangladesh to minimize fear.

To date, no study probably investigated the mucormycosis and/or fungal infection fear in Bangladesh among health care workers to cope with the emerging public health phenomenon. Therefore, the present study investigates the association between mucormycosis fear and insomnia status and other predictors among Bangladeshi healthcare workers during the COVID-19 pandemic. The ongoing pandemic has taught us to be prepared with a capable health system to save lives by being given the top priority of healthcare. Addressing the association between fear of mucormycosis and insomnia among health care workers, including knowledge of mucormycosis, sociodemographic, work-related, COVID-19, and health-related information, may help to take appropriate measures to improve their mental health condition, especially with the emergence of mucormycosis like a fungal infection.

## 2. Method

### 2.1. Study Settings

This cross-sectional study was conducted between 27 May 2021 to 03 June 2021 in Bangladesh. The inclusion criteria include (a) the registered health care workers; doctor, nurse, and others (technologist, radiologist, and medical assistant), (b) working in clinical settings during the COVID-19 associated mucormycosis cases were found, (c) willing to participate in this study by an online consent, and (d) responded to all items of the questionnaire.

### 2.2. Data Collection Procedure

Due to lockdown, a face-to-face interview was strictly restrained. Thus, we followed convenience and snowball sampling methods during the pandemic situation. For data collection, a semi-structured online questionnaire (in “Google Form”) was used. The data were collected via available social media platforms. By clicking on the link, the participants viewed the aim and objective of the study on the first page. On the second page, the sociodemographic and workplace-related items were presented. The comorbidity, health, and smoking status-related items were presented on the third page. The items of knowledge measure were presented on the third page. On the last page, items of ISI-3 and mucormycosis fear measure were presented. However, from 468 responses, a total of 422 completed responses were included in the final analysis.

### 2.3. Background Characteristics

The sociodemographic information such as age, gender, residence, profession, marital status, having children, and living status were collected in this study. Work-related information included job type, working condition, having PPE (personal protective equipment), and directed patient care. We included COVID-19 related information of the participants; COVID-19 positive status, friend and family members’ (FNF), COVID-19 positive status, and FNF COVID-19 death status. Additionally, health-related information included comorbidities status, perceived health status, and smoking status.

### 2.4. Comorbidities Status

The comorbidity status of the respondents was assessed by asking whether they have any comorbidities from the given options; no comorbidity, asthma, diabetes, hypertension, heart disease, cardiovascular disease, chronic kidney disease, chronic liver disease, cancer, and others. The comorbidity status was considered as “no” when the response was “no comorbidity,” and the rest of the responses were considered as comorbidity status “yes.”

### 2.5. Perceived Health Status

The World Health Organization (WHO) recommended a single-item scale (“In general, how would you rate your current health status?”) that was used to measure the perceived health status in this [32]. The responses of the scale were at five options “very good,” “good,” “fair,” “bad,” or “very bad.” For statistical analysis, “very good” and “good” were considered as “healthy,” whereas “fair” was as “relatively healthy” [33]. Similarly, “bad” and “very bad” were considered as “unhealthy.”

### 2.6. Knowledge Score of Mucormycosis

The knowledge scores were accessed by authors’ adopted six items measure based on a five-point Likert scale; 1 for strongly disagree to 5 for strongly agree. The overall score was obtained by summing all the items, whereas the scores range from 6 to 30. A higher score indicated a higher level of knowledge. The measure is given in **supplementary file-1**. Experts’ opinions were obtained for the content validity of the measure. Some modifications were performed based on their suggestions. Inter items correlations were accessed for the construct validity of the measure. The items were found significantly correlated. However, the physicians’ mucormycosis knowledge score was found highest, which was expected. The reliability coefficient of the measure was found on an acceptable range (Cronbach’s α = 0.79).

### 2.7. Insomnia Status

Three items’ Insomnia Severity Index-3 (ISI-3), were used to assess the respondents’ insomnia status in the past 2 weeks [34]. The responses of this brief screening tool items were at five points Likert scale, and the total score ranges from 0 to 12. A score ≥ 7 considers insomnia status ‘yes’ and <7 as ‘no.’ However, the reliability coefficient of ISI-3 (Cronbach’s α = 0.87) showed excellent internal consistency in this study.

### 2.8. Mucormycosis Fear Score

A nine items measure was used to assess fear associated with the mucormycosis (presented in **Table 1**) that was inspired by Wheaton et al. and Blakey et al., who assessed the H1N1 and Ebola-related fear respectively [35], [36]. The items were responded from 0 (not at all) to 4 (very much), where the sum score ranged from 0 to 36. The higher score indicated a higher level of mucormycosis fear. In this study, the nine items measure demonstrated good internal consistency with an overall reliability coefficient of 0.82. After deletion of each item, none of the reliability coefficients was found less than 0.78.

**Table 1:**
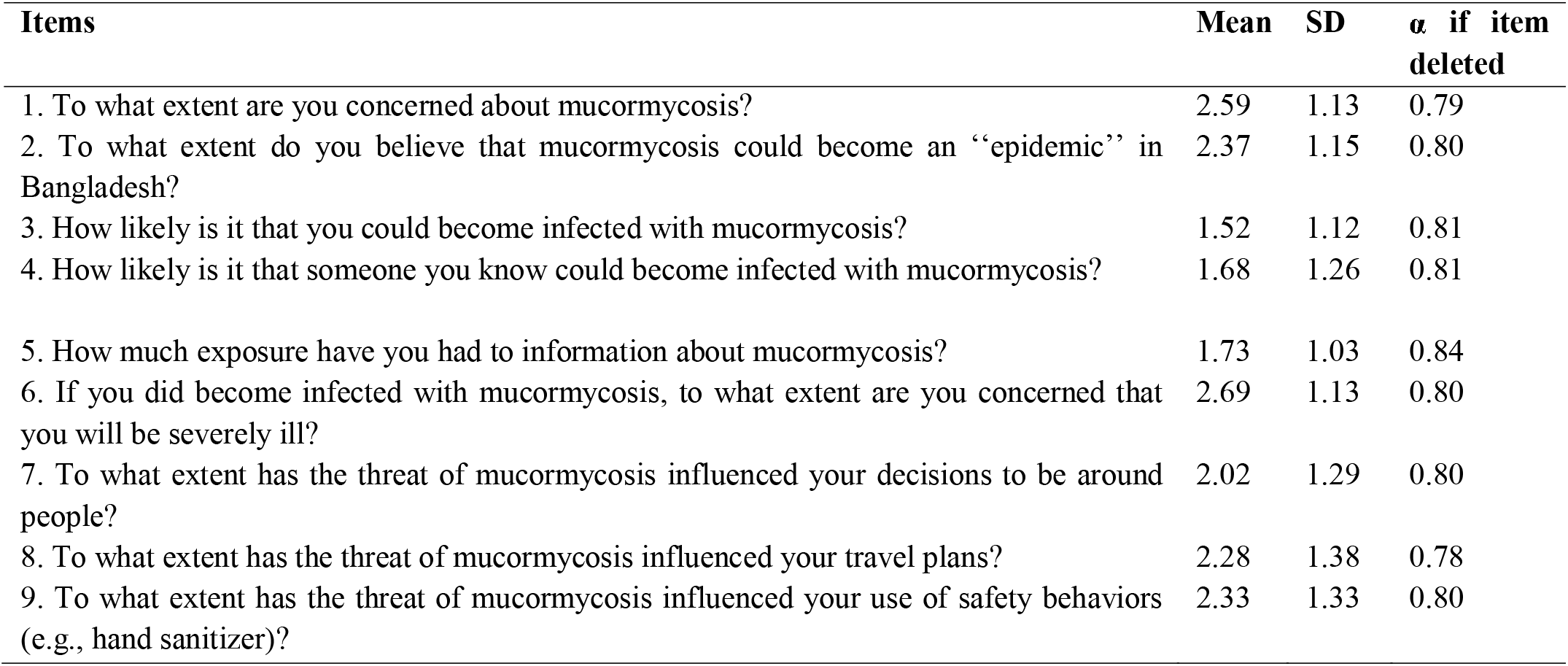
Item Properties of the Mucormycosis Measure.

| Items | Mean | SD | $\alpha$ if item deleted |
| --- | --- | --- | --- |
| 1. To what extent are you concerned about mucormycosis? | 2.59 | 1.13 | 0.79 |
| 2. To what extent do you believe that mucormycosis could become an “epidemic” in Bangladesh? | 2.37 | 1.15 | 0.80 |
| 3. How likely is it that you could become infected with mucormycosis? | 1.52 | 1.12 | 0.81 |
| 4. How likely is it that someone you know could become infected with mucormycosis? | 1.68 | 1.26 | 0.81 |
| 5. How much exposure have you had to information about mucormycosis? | 1.73 | 1.03 | 0.84 |
| 6. If you did become infected with mucormycosis, to what extent are you concerned that you will be severely ill? | 2.69 | 1.13 | 0.80 |
| 7. To what extent has the threat of mucormycosis influenced your decisions to be around people? | 2.02 | 1.29 | 0.80 |
| 8. To what extent has the threat of mucormycosis influenced your travel plans? | 2.28 | 1.38 | 0.78 |
| 9. To what extent has the threat of mucormycosis influenced your use of safety behaviors (e.g., hand sanitizer)? | 2.33 | 1.33 | 0.80 |

### 2.9. Data Analysis

Descriptive statistics were performed for the background characteristics of the study participants. The overall reliability coefficient of nine items’ fear measure was calculated along with reliability coefficients after deletion of each item. A multivariable linear regression model was fitted to find the association between mucormycosis fear and knowledge of mucormycosis and with the background characteristics. Four hierarchical regression models were fitted to investigate the contributory role of the studied variables on healthcare workers’ mucormycosis fear. The p-value <0.05 was considered statistically significant at a 95% confident interval. Data were analyzed by using statistical software STATA-16.

## 3. Result

### 3.1. Characteristics of the Study Population

In this study, 422 Bangladeshi healthcare workers participated. In **Table 2**, the background characteristics are presented. One-third of the participants (30.57%) was found the presence of insomnia. The mean score of knowledge was 24.67 (SD: 4.57). The majority of them (82.94%) were aged between 24 years to 30 years. Female participants were maximum (52.37%). Most of them were from urban (88.15%) areas. In terms of religion, most of them were Muslim (80.57%). In this study, 45.26% were doctors, 38.15% were nurses, and 16.59% were other healthcare workers. The majority of them were married (54.27%), and 25.12% had children. However, 21.09% of them lived alone. Almost half of them did private jobs (51.42%). One-third of them worked in COVID-19 condition (31.52%), and the majority of them (65.17%) had PPE. More than half of them (74.88%) were involved with direct patient care. One-third of the respondents (27.96%) got COVID-19 positive status, and more than two-thirds (75.53%) of their FNF got COVID-19 positive status. Even 22.09% of their FNF died of COVID-19 infection. Having comorbidities status was 20.38%. Interm of perceived health status, more than two-thirds (74.17%) was unhealthy, 20.85% was perceived relatively healthy. Most of them (81.04%) were never smoked.

**Table 2:**
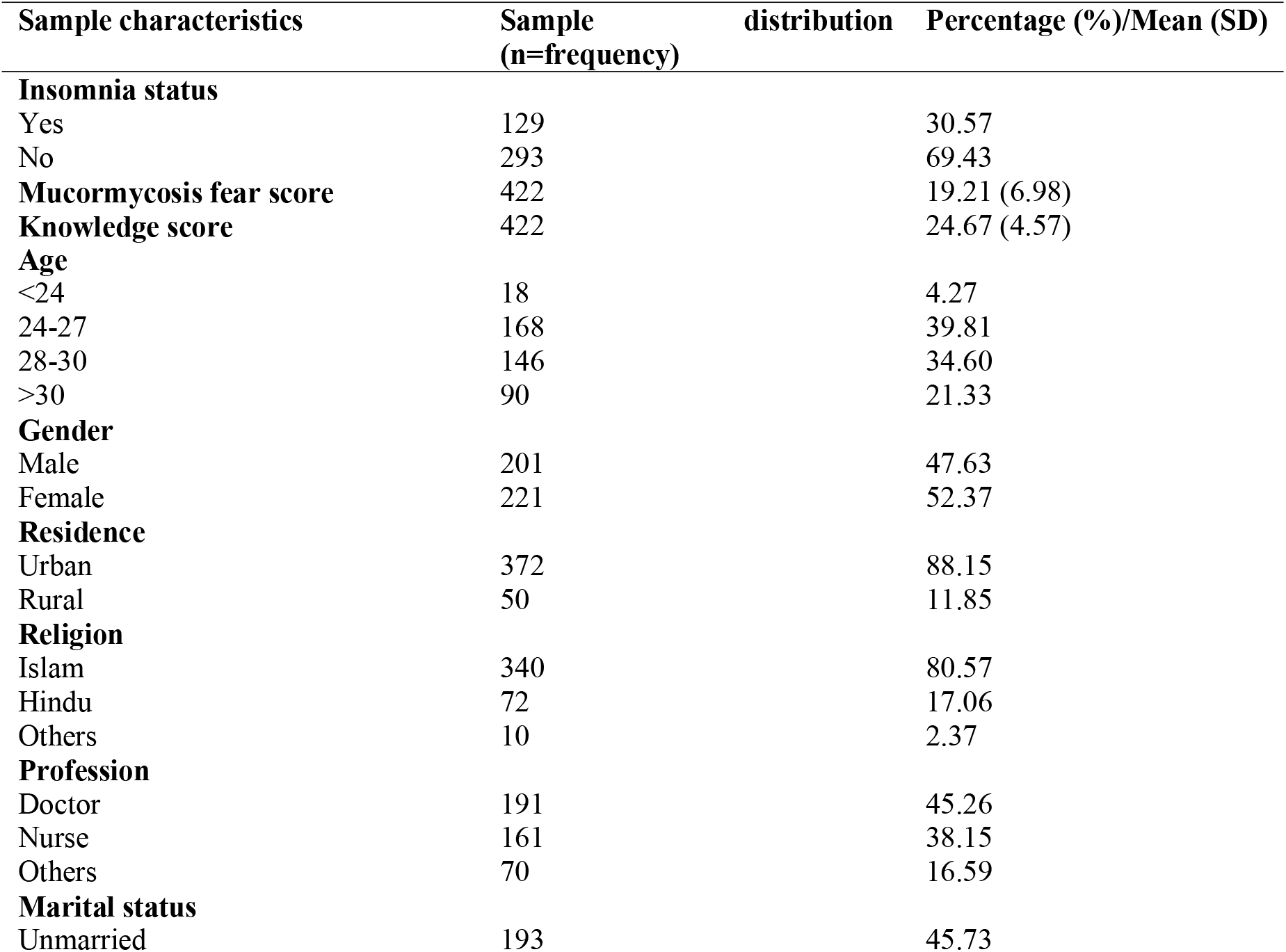
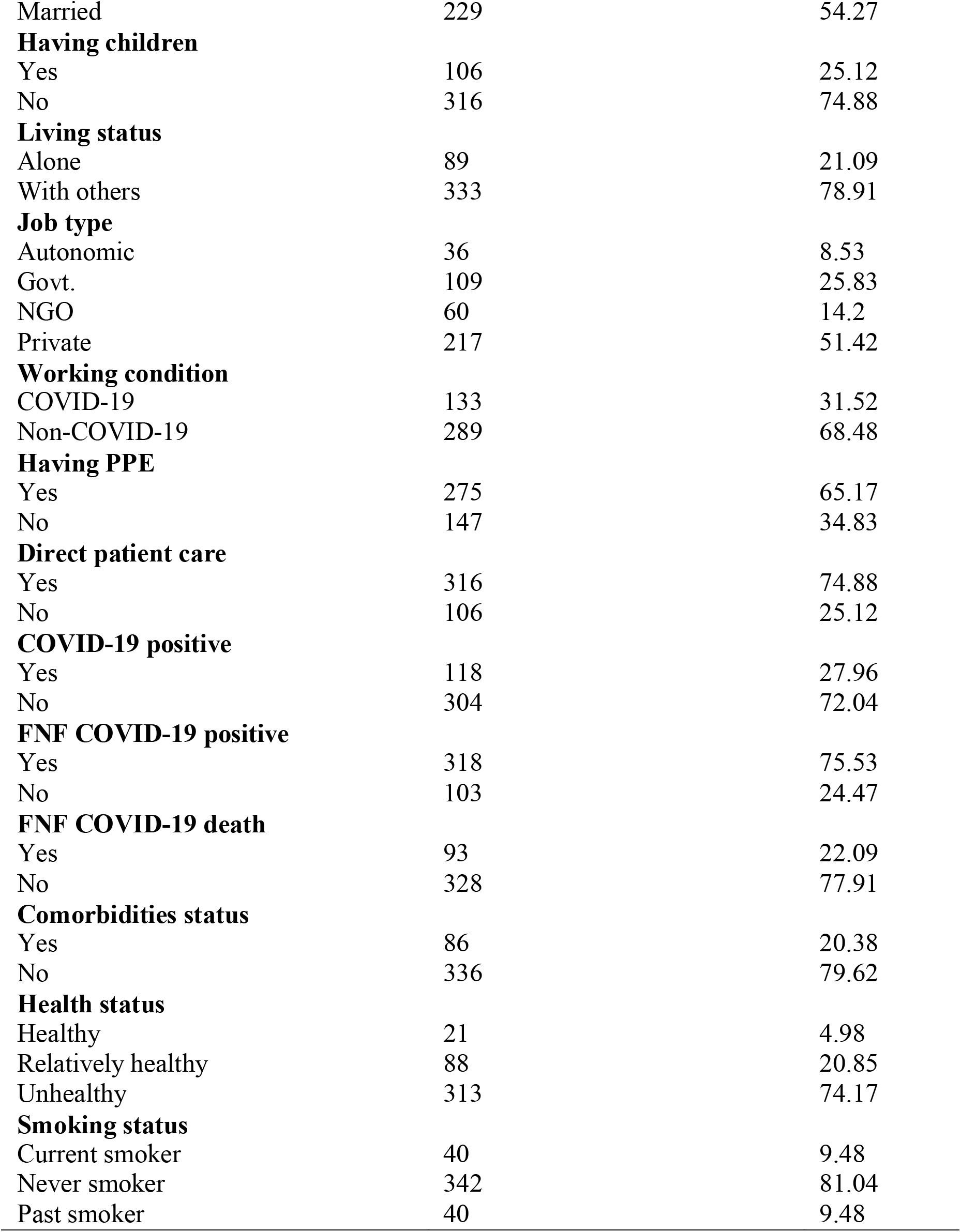
Characteristics of the Study Participants (n=422)

### 3.2. Association between Fear of Mucormycosis and Insomnia Status and other Predictors

Table 3 shows the association between mucormycosis fear and insomnia status and other predictors’ variables. In the adjusted Multivariable Linear Regression model, results showed that respondents with insomnia status had a higher score of mucormycosis fear than not having (β = 3.91, 95% CI: 2.49, 5.33, *p* <0.001). With the increased knowledge score, the average score of mucormycosis fear was significantly increased (β = 0.35, 95% CI: 0.20, 0.50, *p* <0.001). Regarding gender, the fear score was found significantly higher among the females than males (β = 2.12, 95% CI: 0.56, 3.57, *p* = 0.008). However, the fear score was significantly higher among the nurses than the doctors (β = 2.02, 95% CI: 0.14, 3.89, *p* = 0.035). Moreover, the respondents’ FNF who died of COVID-19 had a higher fear score, significantly (β = 2.58, 95% CI: 0.90, 4.26, *p* = 0.003). The rest of the variables, i.e., age, residence, religion, marital status, having children, living status, job type, working condition, having PPE, direct patient care, COVID-19 positive, FNF COVID-19 positive, comorbidities status, health status, smoking status showed no significant association with the mucormycosis fear score in the adjusted model.

### 3.3. Predictive Models of Health Care Workers’ Mucormycosis Fear

The hierarchical regression models are presented in **Table 4**. The four models were fitted; wherein model 1, only insomnia status and the knowledge score were included. The sociodemographic information was included in model 2, and the work-related information was included in model 3. However, COVID-19 and health-related information were added in model 4. All of the models were found to predict the health care workers’ mucormycosis fear significantly. The insomnia status and knowledge score of mucormycosis explained almost 12.20% variance of mucormycosis fear. With the inclusion of demographic information, the total explained variance was increased by 16.20%. After adding the worked-related information, the explained variance was not altered. Moreover, the inclusion of COVID-19 and health-related information in the 4^th^ model, 2% explained variance was increased, and the final model explained a total 18.20% variance of the mucormycosis fear.

**Table 3:** Association between Fear of Mucormycosis and Insomnia Status and Other Predictors (n=422)

| Sample Characteristics | Unadjusted $\beta$ (95% CI) | *p*-value | Adjusted $\beta$ (95% CI) | *p*-value |
| --- | --- | --- | --- | --- |
| **Insomnia status** |  |  |  |  |
| Yes | 3.94 (2.53, 5.34) | **<0.001** | 3.91 (2.49, 5.33) | **<0.001** |
| No | Reference |  | Reference |  |
| **Knowledge score** | 0.36 (0.22, 0.50) | **<0.001** | 0.35 (0.20, 0.50) | **<0.001** |
| **Age** |  |  |  |  |
| <24 | Reference |  | Reference |  |
| 24-27 | -1.59 (-5.00, 1.82) | 0.359 | -2.31 (-5.61, 0.99) | 0.169 |
| 28-30 | -1.56 (-4.99, 1.88) | 0.374 | -2.03 (-5.52, 1.46) | 0.254 |
| >30 | -0.80 (-4.35, 2.75) | 0.658 | -1.87 (-5.66, 1.92) | 0.332 |
| **Gender** |  |  |  |  |
| Male | Reference |  | Reference |  |
| Female | 2.78 (1.47, 4.10) | **<0.001** | 2.12 (0.56, 3.67) | **0.008** |
| **Residence** |  |  |  |  |
| Urban | 1.97 (-0.10, 4.03) | 0.061 | 1.63 (-0.42, 3.68) | 0.119 |
| Rural | Reference |  | Reference |  |
| **Religion** |  |  |  |  |
| Islam | Reference |  | Reference |  |
| Hindu | 1.17 (-0.61, 2.94) | 0.197 | 0.26 (-1.49, 2.01) | 0.771 |
| Others | 4.08 (-0.31, 8.47) | 0.068 | 3.25 (-0.96, 7.45) | 0.130 |
| **Profession** |  |  |  |  |
| Doctor | Reference |  | Reference |  |
| Nurse | 0.48 (-0.99, 1.95) | 0.521 | 2.02 (0.14, 3.89) | **0.035** |
| Others | -1.08 (-3.10, 0.84) | 0.268 | 1.99 (-0.211, 4.19) | 0.076 |
| **Marital status** |  |  |  |  |
| Unmarried | Reference |  | Reference |  |
| Married | -0.09 (-1.44, 1.25) | 0.891 | -0.20 (-1.82, 1.42) | 0.805 |
| **Having children** |  |  |  |  |
| Yes | 0.06 (-1.49, 1.60) | 0.944 | 0.32 (-1.51, 2.14) | 0.734 |
| No | Reference |  | Reference |  |
| **Living status** |  |  |  |  |
| Alone | Reference |  | Reference |  |
| With others | 0.53 (-1.11, 2.17) | 0.528 | 0.70 (-.89, 2.30) | 0.346 |
| **Job type** |  |  |  |  |
| Autonomic | Reference |  | Reference |  |
| Govt. | 0.22 (-2.42, 2.85) | 0.873 | -0.32 (-3.05, 2.41) | 0.815 |
| NGO | -1.73 (-4.62, 1.17) | 0.241 | -0.60 (-3.47, 2.28) | 0.684 |
| Private | -0.40 (-2.87, 2.07) | 0.748 | -0.18 (-2.57, 2.21) | 0.881 |
| **Working condition** |  |  |  |  |
| COVID-19 | Reference |  | Reference |  |
| Non-COVID-19 | -0.35 (-1.79, 1.09) | 0.636 | 0.24 (-1.32, 1.81) | 0.761 |
| **Having PPE** |  |  |  |  |
| Yes | 0.94 (-0.46, 2.34) | 0.186 | 0.77 (-.76, 2.30) | 0.325 |
| No | Reference |  | Reference |  |
| **Direct patient care** |  |  |  |  |
| Yes | 0.58 (-0.97, 2.12) | 0.464 | 0.14 (-1.44, 1.71) | 0.864 |
| No | Reference |  | Reference |  |
| **COVID-19 positive** |  |  |  |  |
| Yes | 0.25 (-1.25, 1.74) | 0.747 | -0.16 (-1.66, 1.35) | 0.838 |
| No | Reference |  | Reference |  |
| **FNF COVID-19 positive** |  |  |  |  |
| Yes | -0.672 (-2.23, 0.89) | 0.396 | -1.00 (-2.59, 0.57) | 0.209 |
| No | Reference |  | Reference |  |
| **FNF COVID-19 death** |  |  |  |  |
| Yes | 2.48 (0.89, 4.07) | **0.002** | 2.58 (0.90, 4.26) | **0.003** |
| No | Reference |  | Reference |  |
| **Comorbidities status** |  |  |  |  |
| Yes | 0.13 (-1.53, 1.79) | 0.881 | 0.09 (-1.57, 1.76) | 0.912 |
| No | Reference |  | Reference |  |
| **Health status** |  |  |  |  |
| Healthy | Reference |  | Reference |  |
| Relatively healthy | 0.68 (-2.66, 4.02) | 0.689 | 0.94 (-2.28, 4.16) | 0.566 |
| Unhealthy | 0.03 (-3.07, 3.13) | 0.984 | 1.50 (-1.56, 4.57) | 0.336 |
| **Smoking status** |  |  |  |  |
| Current smoker | Reference |  | Reference |  |
| Never smoker | 1.21 (-1.09, 3.50) | 0.301 | -0.83 (-3.25, 1.58) | 0.499 |
| Past smoker | 0.35 (-2.72, 3.42) | 0.823 | -0.09 (-3.06, 2.88) | 0.953 |

**Table 4:** Predictive Models of Mucormycosis Fear (n=422)

|  | **Model 1** (F = 29.01, R<sup>2</sup> = 0.122, adjusted R<sup>2</sup> = 0.118, p <0.001) |  | **Model 2** (F =7.92, R<sup>2</sup> = 0.162, adjusted R<sup>2</sup> = 0.141, p <0.001) |  | **Model 3** (F = 5.72, R<sup>2</sup> =0.164, adjusted R<sup>2</sup> = 0.136, p <0.001) |  | **Model 4** (F =4.44, R<sup>2</sup> =0.182, adjusted R<sup>2</sup> =0.141, p <0.001) |  |
| --- | --- | --- | --- | --- | --- | --- | --- | --- |
| **Variables** | **$\beta$** | **95% CI** | **$\beta$** | **95% CI** | **$\beta$** | **95% CI** | **$\beta$** | **95% CI** |
| Insomnia status<sup>a</sup> | -3.90 | -5.26, -2.54 | -3.82 | -5.18, -2.46 | -3.87 | -5.24, -2.50 | -3.99 | -5.39, -2.58 |
| Knowledge score | 0.36 | 0.22, 0.49 | 0.36 | 0.22, 0.51 | 0.36 | 0.21, 0.50 | 0.34 | 0.19, 0.48 |
| Age<sup>b</sup> |  |  | -0.06 | -0.94, 0.82 | -0.10 | -1.00, 0.81 | -0.23 | -1.16, 0.70 |
| Gender<sup>c</sup> |  |  | -2.05 | -3.35, -0.76 | -1.98 | -3.29, -0.66 | -1.83 | -3.15, -0.51 |
| Residence<sup>d</sup> |  |  | 1.67 | -0.28, 3.61 | 1.66 | -0.31, 3.62 | 1.66 | -0.32, 3.64 |
| Religion<sup>e</sup> |  |  | 0.75 | -0.63, 2.13 | 0.72 | -0.66, 2.11 | 0.93 | -0.46, 2.33 |
| Profession<sup>f</sup> |  |  | 0.79 | -0.17, 1.75 | 0.71 | -0.28, 1.69 | 0.87 | -0.16, 1.90 |
| Marital status<sup>g</sup> |  |  | -0.44 | -2.00, 1.12 | -0.40 | -1.97, 1.16 | -0.35 | -1.92, 1.21 |
| Having children<sup>h</sup> |  |  | 0.44 | -1.29, 2.17 | 0.50 | -1.25, 2.24 | 0.57 | -1.17, 2.32 |
| Living status<sup>i</sup> |  |  | 0.82 | -0.74, 2.38 | 0.85 | -0.72, 2.42 | 0.88 | -0.70, 2.46 |
| Job type<sup>j</sup> |  |  |  |  | -0.03 | -0.65, 0.59 | -0.11 | -0.74, 0.51 |
| Working condition<sup>k</sup> |  |  |  |  | 0.24 | -1.29, 1.77 | 0.11 | -1.42, 1.64 |
| Having PPE<sup>l</sup> |  |  |  |  | 0.77 | -0.70, 2.24 | 0.57 | -0.93, 2.06 |
| Direct patient care<sup>m</sup> |  |  |  |  | 0.33 | -1.18, 1.86 | 0.23 | -1.29, 1.76 |
| COVID-19 positive<sup>n</sup> |  |  |  |  |  |  | -0.23 | -1.71, 1.26 |
| FNF COVID-19 positive<sup>o</sup> |  |  |  |  |  |  | -0.96 | -2.52, 0.61 |
| FNF COVID-19 death<sup>p</sup> |  |  |  |  |  |  | 2.40 | 0.76, 4.04 |
| Comorbidities status<sup>q</sup> |  |  |  |  |  |  | 0.21 | -1.42, 1.83 |
| Health status<sup>r</sup> |  |  |  |  |  |  | 0.65 | -0.54, 1.84 |
| Smoking status<sup>s</sup> |  |  |  |  |  |  | -0.01 | -1.46, 1.48 |

a, h, l, m, n, o, p 1= Yes, 2= No. b 1= <25, 2= 24-27, 3= 28-30, 4= >30. c 1= Male, 2= Female, d 1= Urban, 2= Rural. e 1= Islam, 2= Hindu, 3= Others. f 1= Doctor, 2= Nurse, 3= Others. g 1= Unmarried, 2= Married. i 1= Alone, 2= With others. j 1= Autonomic, 2= Govt., 3= NGO, 4= Private. k 1= COVID-19, 2= Non-COVID-19. q 1= Healthy, 2= Relatively healthy, 3= Unhealthy. r 1= Current smoker, 2= Never smoker, 3= Past smoker.

## 4. Discussion

This study found that mucormycosis fear is significantly associated with the insomnia status among Bangladeshi health care workers. Alongside, numerous predictors like knowledge of mucormycosis, age, gender, profession, and FNF died of COVID-19 were found significantly associated with mucormycosis fear. Albeit, the fear of infectious diseases during the outbreak has been recorded previously in many studies in various aspects [37]–[40].

The association between mucormycosis fear and insomnia status was explored. This finding can be explained by Cawcutt et al., who found a strong association between insomnia and COVID-19 fear among health workers [41]. Compared to the non-exposed, exposure to COVID-19 showed to alter the healthcare workers’ quality of sleep [42]. Few Italian studies showed that the fear of COVID-19 and insomnia were more significant in the first pandemic wave of the pandemic [43], and in the second wave, fear turned to stress [44], and finally, in the third wave, sleep slightly improved [45]. In Bangladesh, M. Hossain et al. also reported that during the COVID-19 pandemic, sleep disturbance had been increased [46]. Therefore, health care workers having insomnia may become prone to mucormycosis fear during COVID-19.

In this study, mucormycosis fear and knowledge were found significantly associated in both unadjusted and adjusted linear models. With the increase of knowledge score, the mucormycosis fear score was significantly increased. Similarly, Govindharaj et al. reported that knowledge of the disease and fear were strongly associated with each other [47]. An earlier study reported that the mortality rate due to mucormycosis was 33%, but it has been reported much higher in this COVID-19 era [48]. Alongside, multiple crucial steps for removing the organs have been reported for the mucormycosis management procedure [15]. Thus, based on the study findings, increment of knowledge of mucormycosis regarding the severity of mucormycosis for COVID-19 infection may facilitate mucormycosis fear.

The senior age groups were found to be less fearful of mucormycosis compared to the junior age group, though age was found statistically insignificant in this study. This study found that mucormycosis fear was significantly associated with female health workers compared to males. A study revealed that fear is more prevalent in females than males in the COVID-19 pandemic in Bangladesh [49].

We found a significant association of mucormycosis fear with the nursing profession compared with other healthcare professionals. As nurses are involved, they are more prone to direct contact-related care, which may induce more fear among them than other health workers. This working nature of the nurses may propel more fear. The Bangladeshi nurses had mental health-related symptoms during the first web of the COVID-19 pandemic [50]. Similarly, Tayyib et al. reported a high level of fear and stress during the COVID-19 context [51].

A significant relationship was found between the fear of mucormycosis and COVID-19 death incidence of family and friends. Family and friends are the two most important aspects of one’s life. Naturally, one related to them thinks about their wellbeing and other aspects [46]. Again, thinking is derived from the information stored in the memory [52]. Consequently, the death memory of friends or family members was found to be associated with fearfulness.

Mucormycosis fear was found high among the comorbid health workers, though it was not statistically significant. Syndrome et al. reported in a systematic review that having comorbidity is an independent risk factor of mucormycosis infection [7]. Therefore, the comorbid health workers may perceive more mucormycosis fear.

Based on this study finding, the healthcare workers are fearsome toward mucormycosis. The COVID-19, along with mucormycosis cases risen, might pose synergistic fear among the healthcare workers as they confront more risk. Therefore, our study recommends that the mental health of healthcare workers should be more prioritized. Arranging necessitous programs, training, and workshops to cope with the situation can also be commenced.

## 5. Strength and Limitation

To our knowledge, this is the first study in Bangladesh that focused on assessing the association between mucormycosis fear and insomnia status among the health care workers so far, which is a strength of this study. The findings of this study can help the policymakers to improve strategic plans towards COVID-19 associated mucormycosis and/or infectious/fungal disease emergences in a developing country like Bangladesh.

As the nature of the cross-sectional study, causality between the variables could not explain. However, following convenient and snowball sampling methods might have some selection bias. Hence, it is recommended for further researches to assess mucormycosis fear on a larger scale and the sample size among the health care workers of Bangladesh to strengthen the health care system during the pandemic.

## 6. Conclusion

Fungal infection and mucormycosis are not new phenomena, but the emergence of mucormycosis associated with COVID-19 is a big concern. The healthcare workers were playing the most pivotal role in ameliorating the COVID-19 catastrophe. Therefore, the growing concern of mucormycosis cases in Bangladesh during COVID-19 may induce fear among the health care workers. This study documented that nurses were significantly more fearful compared to doctors. The mucormycosis fear was significantly associated with health care workers’ insomnia status, knowledge of mucormycosis, gender, profession, and a friend or family members’ COVID-19 death. The outcomes of this study may help policymakers establish a proactive mental health support system for the health care workers not only for mucormycosis but also for this particular infectious emerging concern. These might be bona fide essential to create an ecosystem from the side of healthcare workers toward patients in need of the highest health support.

## Data Availability

Data will be available by requesting the corresponding author through email.

## Ethics approval and consent to participate

The Institutional Review Board (IRB) of North South University, Bangladesh approved (IRB: 2021/OR-NSU/IRB/0603). The aim and objective were explained on the first page of the survey questionnaire. Those who were willing to participate by online consent were considered as respondents of this study.

## Consent for publication

Not applicable.

## Availability of Data Materials

Dataset used in this study will be available as per request (mailing to the corresponding author).

## Author Contributions

Conceptualization, M.K.H., H.K.; methodology, M.K.H., H.K.; validation and scrutinization, D.K.M, M.K.H., H.K.; investigation, M.K.H., H.K.; writing-original draft preparation, M.K.H. H.K., M.R., A.K.R., S.A.; review and editing, D.K.M., M.K.H., H.K.; supervision, D.K.M.; All authors have read and agreed to the current version of the manuscript.

## Acknowledgments

We would like to accolade all the research assistants of the project who assisted in data collection.

## Tables

**Supplementary Table:**
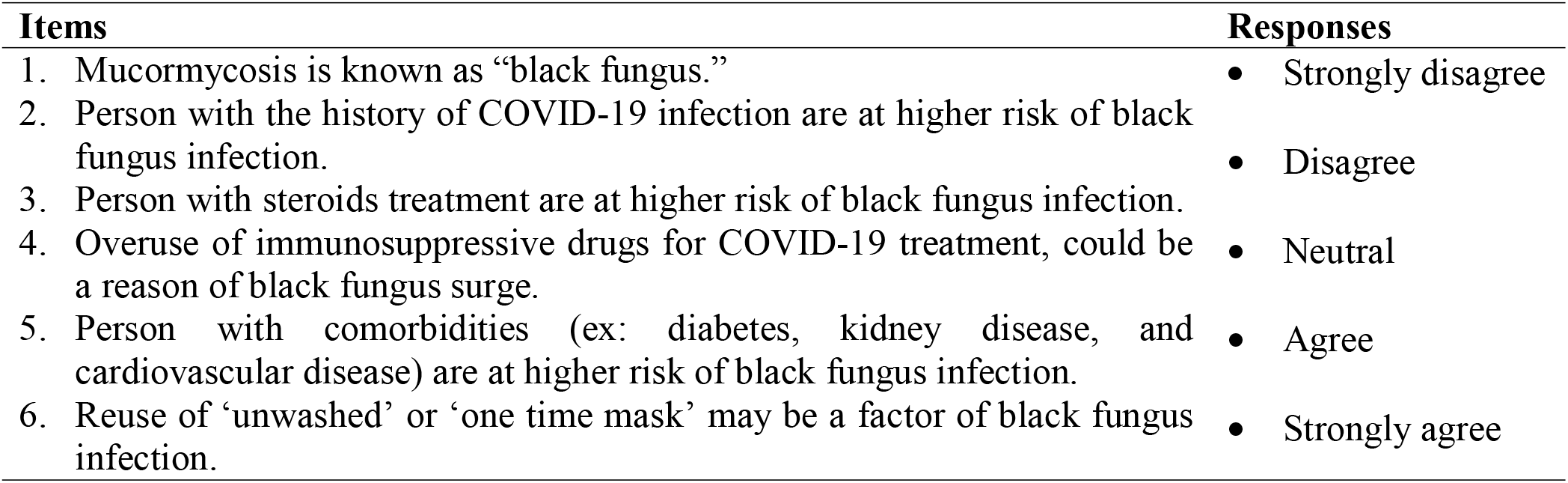
The items of knowledge of mucormycosis (n = 422)

