## Supplementary file_MS ID #259670 for "Predictors of black fungus fear during the COVID-19 pandemic among the Bangladeshi health workers: a cross-sectional study" (from another version of this preprint: version 1, 2021-07-07)

Supplementary Table: The Items of Perception and Attitude Regarding Black Fungus (n = 422)

| Items | Options |
| --- | --- |
| 1. Mucormycosis is known as “black fungus” | - Strongly disagree      - Disagree - Neutral - Agree - Strongly agree |
| 2. Person with the history of COVID-19 infection are at higher risk of black fungus infection. |  |
| 3. Person with steroids treatment are at higher risk of black fungus infection |  |
| 4. Overuse of immunosuppressive drugs for COVID-19 treatment, could be a reason of black fungus surge |  |
| 5. Person with comorbidities (ex: diabetes, kidney disease, cardiovascular disease) are at higher risk of black fungus infection |  |
| 6. Reuse of ‘unwashed’ or ‘one time mask’ may be a factor of black fungus infection |  |
